# The Influence of Coronavirus Disease-2019 (COVID-19) On Parkinson’s Disease: An Updated Systematic Review

**DOI:** 10.1101/2021.05.09.21256929

**Authors:** Vikash Jaiswal, Danah Alquraish, Zouina Sarfraz, Shavy Nagpal, Prakriti Singh Shrestha, Dattatreya Mukherjee, Prathima Guntipalli, Azza Sarfraz, Diana F Sánchez Velazco, Arushee Bhatnagar, Saloni Savani, Elmjedina Halilaj, Samir Ruxmohan, Wilson Cueva

## Abstract

**Background:** COVID-19 has affected global communities with multiple neurological complications in addition to other critical medical issues. COVID-19 binds to the host’s angiotensin-converting enzyme2 (ACE2) receptors, which are expressed in the neurons and glial cells, acting as an entry port to the central nervous system (CNS). ACE2 receptors are abundantly expressed on dopamine neurons, which may worsen the prognosis of motor symptoms in Parkinson’s disease (PD). SARS-CoV-2 may lead to an indirect response via immune-mediated cytokine storms and propagate through the CNS leading to damage. PD is also been associated with certain post-viral infections apart from COVID-19, such as HSV, Influenzavirus A, Measles virus, Cytomegalovirus, and Mumps.

**Objective:** In this systematic review, we aim to provide a thorough analysis of associations between COVID-19 and neurological outcomes for patients with PD.

**Methods:** Using PRISMA statement 2020, a systematic review was conducted to isolate confirmed COVID-19 patients and analyze the PD-associated neurological outcomes. A systematic literature search was conducted using the following databases: PubMed, Science Direct, Google Scholar, and Cochrane databases. The following keywords were used “COVID19, SARS-CoV-2, Parkinson’s disease, Pandemic, Mortality.” A modified Delphi process was employed to include the studies and ensure that the clinical outcome measures were addressed.

**Results:** Of the 355 records located during the initial round of screening, 16 were included in the final synthesis. Of PD patients who tested positive for SARS-CoV-2, worsening motor symptoms and other viral-associated symptoms were reported. These symptoms included bradykinesia, tremors, gait disturbances, delirium and dementia, and severe spasms of arms and legs. Encephalopathy was presented in two of the included studies. Increased mortality rates were identified for hospitalized patients due to COVID-19 and PD as compared to other patient groups, albeit with limited generalizability due to high bias of included studies.

**Conclusion:** Patients with PD may experience substantial worsening of motor and non-motor symptoms due to COVID 19. Given the novelty of neurological-viral associations, clinical studies in the future ought to explore the disease severity and neurological outcomes in COVID-19 positive patients with PD as compared to non-PD patients, in addition to understanding the role of ACE2 in increased vulnerability to contracting the infection and as a treatment modality.

## 1. Introduction

At the onset of the coronavirus disease (2019) pandemic on 31 December 2019, many reports found that SARS-CoV-2 primarily caused pneumonia-like symptoms ^1^. This virus emerged as a global threat and a public health emergency of imminent concern across countries worldwide, with exponential transmission capability. As of May 25, 2021, there have been 167 million confirmed cases of COVID-19, with reported deaths up to 3.47 million globally ^2^. COVID-19 belongs to a novel member of the coronaviridae family named SARS-CoV-2 (Severe Acute Respiratory Syndrome Coronavirus 2) ^3^. Coronaviruses are enveloped positive single-stranded RNA viruses, and the 3’ terminal contains structural proteins. The Spike (S) protein allows the virus to fuse to host cell membranes in proximity to infected and uninfected cells ^4^. A surge in the levels of cytokines (TNF□α, IL□6, IL□8, and IL□10) have been reported, leading to a suppression of T-cell response ^5^. To date, a multitude of studies have assessed the role of angiotensin-converting enzyme2 (ACE2) receptors in increasing vulnerability of hosts, however, there is a dearth of analysis on the role of ACE inhibitors in treating Parkinson’s disease (PD) patients. In this systematic review, we aim to provide an updated, thorough analysis of associations between COVID-19 and neurological outcomes for patients with PD.

Coronavirus disease (2019) has prompted many challenges in global healthcare systems due to its unpredictability and unique manifestations in 1) pulmonary, 2) neurological, 3) cardiovascular, 4) gastrointestinal, and 5) hematological systems. The scientific literature confirms the presence of SARS-CoV-2 in CSF, in addition to respiratory, fecal, and blood samples. In about 36.4% of cases, neurological findings have been identified, ranging from dizziness, headache, hyposmia, hypogeusia, dysphagia, muscle pain, seizures, and loss of consciousness ^6^. COVID-19 also influences socializing factors such as the surge in isolation-induced neurological effects, limited social activities possibly leading to short or long-term neuropsychiatric disorders.

We hypothesize that COVID-19 induced lockdowns, self-isolation, and the fears associated with contracting disease accentuate neuro-psychiatric problems such as depression and cognition. **Figure 1** illustrates motor and non-motor manifestations in addition to autonomic manifestations that are pertinent in the assessment of the clinical outcomes of confirmed positive PD patients. Given the dearth of data, we aim to provide a comprehensive and updated systematic review to guide clinical care and practice in understanding the effects of COVID-19 on Parkinson’s disease.

**Figure 1.**
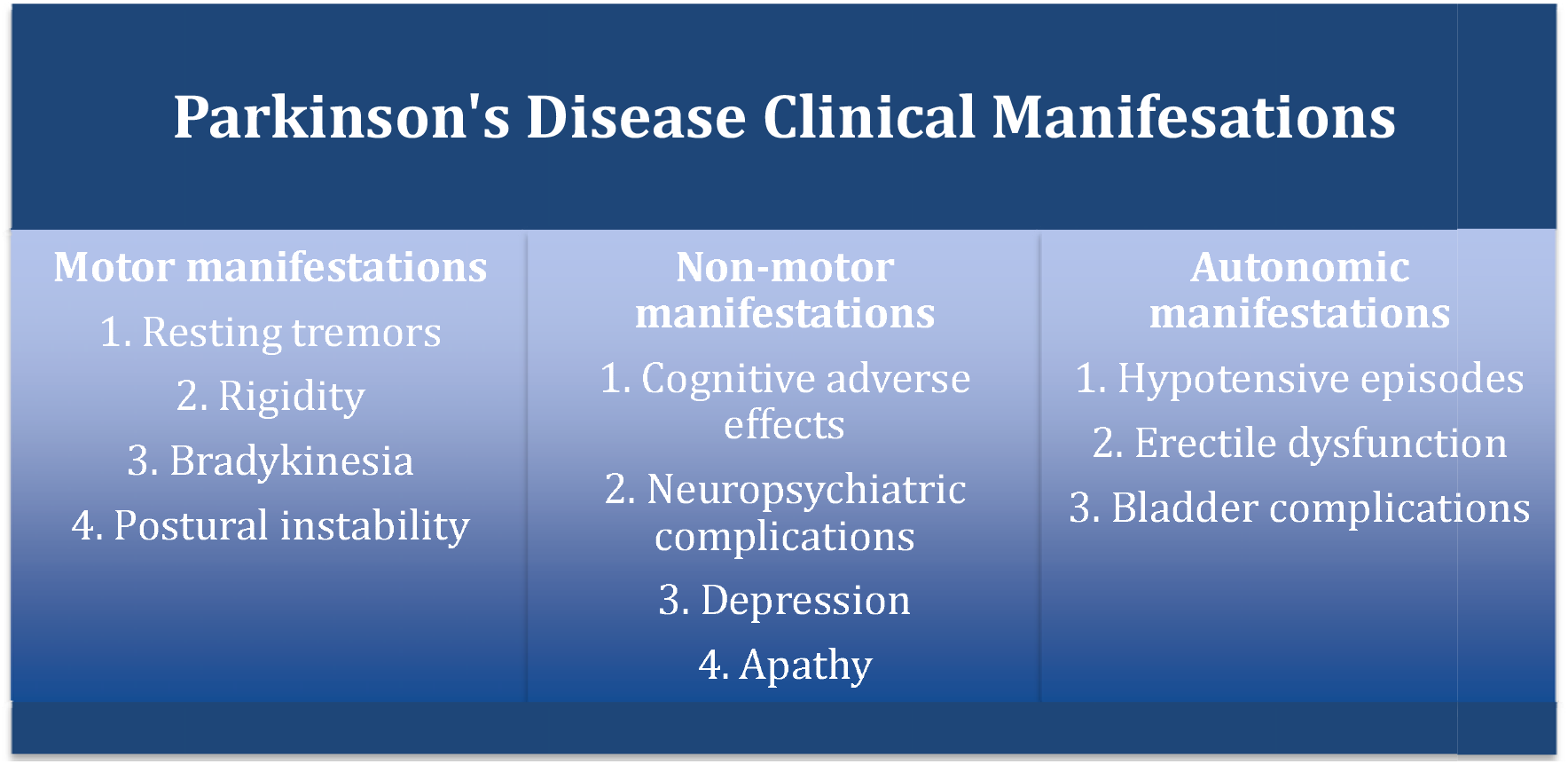
Clinical manifestations of Parkinson’s disease.

## 2. Methodology

### 2.1. Search strategy

Using the Preferred Reporting Items for Systematic Reviews and Meta-Analyses (PRISMA) 2020 statement, a systematic literature search was performed from December 2019 through May 2021 ^7^. The following set of keywords were used, employing the Boolean (and/or) logic: “COVID19”, “SARS-CoV-2”, “Parkinson’s disease,” “Pandemic,” “Mortality.” We searched the following databases, namely, PubMed, Cochrane, Science Direct, EMBASE, and Google Scholar. A modified Delphi approach was utilized to include studies and ensure that the clinical outcome measures are identified in a systematic and unanimous order in the included studies ^8^. The PRISMA flowchart is illustrated in **Figure 2**.

**Figure 2.**
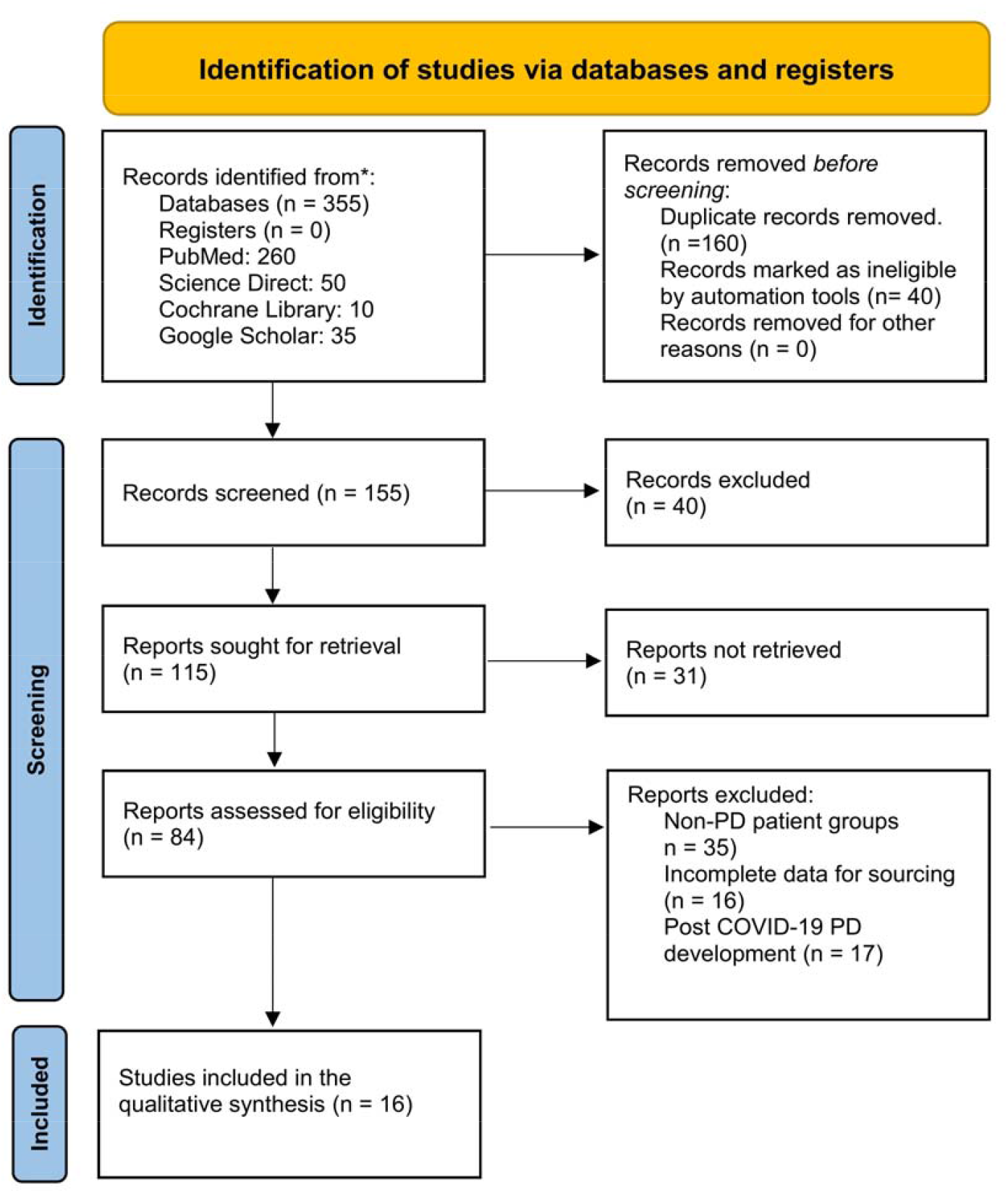
PRISMA Flowchart.

### 2.2. Study selection

We included studies with confirmed COVID-19 disease who had a history of Parkinson’s disease. Only studies published in English and with open access availability were included. In total, 355 records were located during the initial round of screening. During this round of screening, all authors scanned the abstracts and titles independently, which led to the location of 155 records. In the second screening round, full-text records were screened, and 125 articles that met the exclusion criteria were removed. During the third screening round, 84 records were screened for eligibility. In total, 35 records were excluded as non-PD patient groups were present, 16 records had incomplete data for sourcing, and 16 of the full-text records present with post-COVID-19 PD development, which met the exclusion criteria. Finally, 16 articles were included in the qualitative synthesis.

### 2.3. Eligibility criteria

The inclusion criteria for studies and the target population consisted of 1) COVID-19 patients with Parkinson’s disease, 2) patients diagnosed with Parkinson’s disease before COVID-19 infection, 3) age group > 21 years old, 4) male and/or female, and 5) available articles in English language only. Studies were excluded if they met one or more of the following criteria 1) -COVID-19 negative PD patients, 2) studies on pregnant women, 3) the pediatric population, 4) crossover study design, 5) commentaries/perspective pieces.

### 2.4. Data extraction and synthesis

The following data were extracted from eligible studies: serial number, author and year, study design, sample size, gender, age, comorbidities, disease duration, PD outcomes, COVID-19 associated symptoms. A shared spreadsheet was used to input data, which was extracted independently by three authors for analysis and cumulative result interpretation (V.J., D.A., S.N.). The fourth reviewer (Z.S.) solved any discrepancies in data extraction and reach a consensus in case of any disagreements.

Quality assessment was conducted by two authors (V.J. and Z.S.), who independently assessed the (1) criteria of diagnosis of COVID-19 patients, (2) confirmed diagnosis of PD before the COVID-19 pandemic, (3) PD sequelae in the included studies. A descriptive analysis was conducted and presented for the included studies. Percentages and means were presented. Frequent outcomes were reported if they have a frequency of at least 5% or more in the included studies. The term PD complications were based on the motor, non-motor, and autonomic complications mentioned in **Figure 1**.

### 2.5. Role of Funding

No funding was obtained for this study.

## 3. Results

Of the 355 records identified during the first round of screening, 16 studies comprising, 7 (43.8%) were case reports and case series, 4 (25%) case-control studies, 2 (12.5%) cross-sectional, 2 (12.5%) cohort and 1 (6.25%) retrospective study were added (**Figure 3**) ^9,10,11,12,13,14,15,16,17,18,19,20,21,22,23,24^. In total, a synthesis of 1290 PD patients with COVID-19 and the characteristics of all included studies are presented in **Table 1**.

**Table 1.** Characteristics of included studies.

| No | Author | Study Design | Sample (N) PD | Males, % | Age, Mean | Comorbidities % | Disease duration , mean | Outcomes | PD related symptoms With COVID % |
| --- | --- | --- | --- | --- | --- | --- | --- | --- | --- |
| 1 | Cilia R et al, 2020 | Case-control | 12 cases<br>36 control | 41.7 % cases<br>41.7% controls | 65.5 years cases<br>66.3 years control | 8.3% COPD<br>33.3 HTN<br>8.3% obesity<br>8.3% cardiopathy<br>16.7 % cancer<br>8.3% immuno-compromised | 6.3 years cases<br>6.1 years control | 50% affected by COVID-19 (cases group);<br>0% affected by COVID-19 in control group,<br>8.3% hospitalized | Worsening of motor symptoms |
| 2 | Del Prete, Eleonora, et al.2020 | Case-control | 7 cases<br>14 control | 57.2% cases<br>57.2 % control | 75.7 years cases<br>versus<br>75 years control | Cases:<br>71.5% HTN<br>42.8% diabetes<br>28.5% cardiomyopathy<br>14.2% malignancies | 9.3 years cases<br>8.9 years control | 13 % mortality<br>14% case fatality | 30% worsening |
| 3 | Hainque E<br>et al, 2020 | Case<br>report | 2 | 50 % | 77.5 | HTN | 22y | 100 % | 100%<br>worsening |
| 4 | Lo<br>Monaco<br>MR et al,<br>2020 | Case<br>series | 5 | 66.6 % | 74.2 | 20% Chronic Renal<br>failure<br>20% Diabetes<br>hypertension | NA | 20% case<br>fatality | 60%<br>Worsening |
| 5 | Fasano,<br>Alfonso, et<br>al. 2020 | Case-control<br>survey | 105 | 52.4% | 70.5 | Obesity 18.1 %<br>HTN 41.9%<br>COPD 5.6%<br>Diabetes 7.6%<br>Cancer .9 % | 9.9 y | 17.1%<br>hospitalized<br>5.7 % mortality |  |
| 6 | Filatov,<br>Asia, et<br>al.2021 | Case<br>report | 1 |  | 74 | atrial fibrillation<br>cardioembolic<br>stroke<br>COPD | NA | Icu<br>hospitalization<br>Poor prognosis | Encephalopat<br>hy |
| 7 | Brown<br>EG, et al, | Cross<br>sectiona | 51 | 47 % | 65 | Immunocompromis<br>ed 22% | 0–3y□<br>21 | 9.8%<br>Hospitalized | 55 %<br>worsening |
|  | 2020 | 1 |  |  |  | Heart Disease 20%<br>HTN 25%<br>Lung disease 14% | 3–6y 12<br>6–9y 9<br>>9y 9 | 3.9% ICU<br>2% Ventilator | 18 % new PD<br>symptoms |
| 8 | Kobylecki<br>C et al<br>2020 | Cohort,<br>observat<br>ional | 58 | 62.5 % | 78.3 | Diabetes 8%<br>HTN 38% | 9.5 | 22.4% Mortality<br>rate | 69%<br>Delirium<br>54%<br>Dementia |
| 9 | Lo<br>Monaco<br>MR et al,<br>2021 | Case<br>report | 1 | 0% | 58 | NA | 8 y | NA | severe<br>spasms of<br>arms and<br>legs. |
| 10 | Antonini<br>et al 2020 | case<br>series | 10 | 60 % | 78.3 | Diabetes<br>Orthostatic<br>hypotension<br>CHF,<br>COPD<br>Asthma<br>IHD, CKD,<br>dementia,<br>osteoporosis anxiety disorder | 12.7 y | 40% mortality<br>rate | Worsening<br>of mobility<br>with fall<br>Worsening<br>of motor<br>symptoms<br>Worsening of<br>anxiety |
| 11 | Vignatelli et al, 2020 | cohort | 696 PD<br>184 parkinsonism<br>control<br>8590 | 58.8 & PD<br>57.1 parkinsonism<br>58.2 control | 75 PD 80.5 PS<br>Control 76.0 | PD and PS<br>22.6 %<br>cardiomyopathies<br>14.6 %<br>Cerebrovascular disease<br>8.5 % Chronic pulmonary disease<br>.3 % Liver disease<br>5 % Renal disease<br>10% DM<br>8.5% malignancies | NA | Hospitalization<br>0.6% PD<br>3.3% PS<br>0.7% control<br>Mortality rate<br>35.1% in all group | PD:<br>72.8% Tremor<br>80,4%<br>Bradykinesia<br>81.6%<br>Clinical features at onset<br>unilateral<br>PS:<br>52.5% Tremor<br>89.2 %<br>Bradykinesia<br>52.5%<br>Clinical |

**Table 1.** Characteristics of included studies.
|  |  |  |  |  |  |  |  |  | features at onset unilateral |
| --- | --- | --- | --- | --- | --- | --- | --- | --- | --- |
| 12 | Artusi CA,et al, 2020 | multiple case reports | 8 | 62.5% | 74 | HTN<br>diabetes,<br>depression<br>lung neoplasm<br>Atrial fibrillation | 12.2 y | 75% mortality rate | 50% worsening of PD symptoms |
| 13 | Diego Santos-García et al, | cross-sectional | 15 | 47.1% | 76.3 | 26.7% HTN<br>20% DM<br>64.3% Dyslipidemia<br>13.3% Cardiomyopathy<br>6.7% Valvular cardiomyopathy<br>20% Cardiac Arrhythmia<br>6.7% Cardiac Insufficiency<br>14.3% Pulmonary | NA | 33.3% Hospitalized | 65.7% worsening of symptoms<br>47.7% bradykinesia<br>41.4% sleep problems<br>40.7% rigidity<br>34.5% gait disturbances<br>31.3% |
|  |  |  |  |  |  | disease<br>6.7% smoking |  |  | anxiety<br>28.5% pain<br>28.3%<br>fatigue<br>27.6%<br>Depression<br>20.8% tremor |
| 14 | R Sainz<br>Amo et al.<br>2020 | Case<br>Control | Case:<br>33<br>Control:1<br>72 | 5 59 | Case:75.9<br>Control:<br>73.9 | 36% Dementia | Case: 8.9<br>Control8<br>.5 | 54%<br>hospitalization<br>21% mortality<br>rate | NA |
| 15 | Jing li et<br>al.<br>2020 | Case<br>report | 1 | Female | 85 | HTN<br>Stroke | 6 y |  | worsening<br>motor<br>symptoms.<br>Encephalopat<br>hy |
| 16 | De<br>Marcaida<br>et al 2020 | Retrospective | 36<br>22 PD | 64 % | 74.5 | HTN<br>cardiovascular disease<br>renal disease,<br>diabetes<br>chronic lung<br>disease<br>immunosuppressio<br>n | 16.2 y | 67%<br>hospitalization<br>36% mortality | 75%<br>Alteration mental status<br>19 %<br>worsening of<br>their<br>Movement<br>abnormality<br>31%<br>worsening<br>mobility |

**Figure 3.**
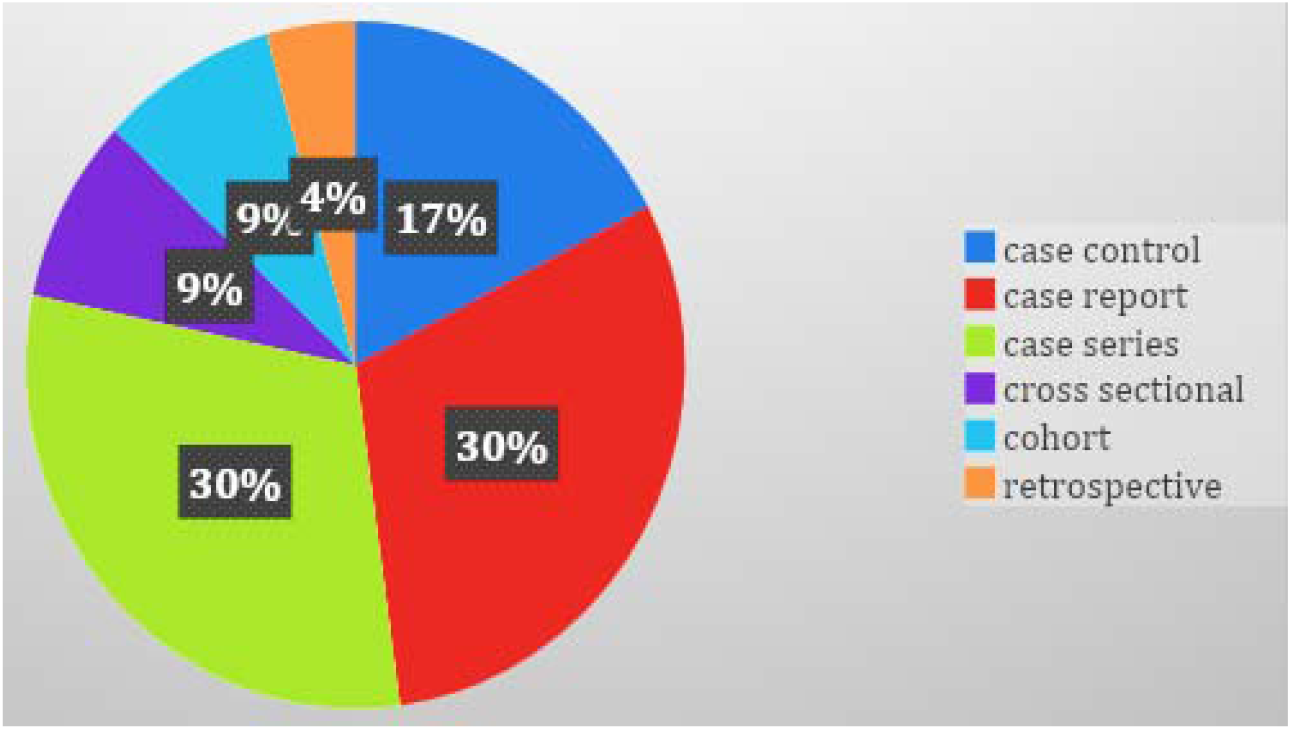
Types of studies included in the systematic review.

Male gender was predominant in 10 studies ^9,10,11,12,13,14,15,16,17,18^ and female gender was the predominant one in 6 studies ^17,19,20,21,22,23^. Of the available patient data, the cumulative mean of age in years of included patients was 76.9 years. The average disease duration spanned 11 years before the incidence of COVID-19 disease. Majority of the included patients presented with comorbid conditions, comprising of hypertension ^9,13,15,16,20,21,23,24^, diabetes ^10,12,16,18,19,20,21^, obesity ^9^, dyslipidemia ^21^, cardiovascular disease ^9,10,14,15,19,20,21,24^, immunocompromised ^9,15,24^, COPD ^9,13,14,18^, asthma ^18^, chronic renal and liver diseases ^18,19,24^. Of the PD patients that were tested and positive infection with SARS-CoV-2, worsening of motor symptoms ^9,10,11,14,15,16,18,19,21,23^ including bradykinesia, tremors, gait disturbances, delirium, and dementia ^13,19^ were noted; severe spasms of the arms and legs ^22^ were found, with individual study percentages of motor symptoms ranging from 19% to 100%, along with other COVD-19 symptoms. Encephalopathy was also one of the major symptoms, which presented in 2 (12.5%) of the 16 studies ^in^ ^20,23^. Mortality rates for PD patients with COVID-19 who were hospitalized ranged from 5.7% to 100% ^9,10,11,12,13,14,15,16,18,19,20,21,22,24^. The mortality was notably 100% due to the inclusion of case reports.

## 4. Discussion

To our best understanding, this is the first systematic review to assess clinical outcomes of Parkinson’s disease (PD) in confirmed COVID-19 patients with a pre-pandemic PD diagnosis. Wide varieties of neurological consequences have been reported in scientific literature among COVID-19 positive patients ^25,26,27,28,29,30,31,32,33^. Neurological symptoms comprised of the following include those associated with core dysfunction (fatigue, headache, confusion, stroke ^34^, dizziness, syncope ^35^, seizure, anorexia, and insomnia) ^36,37,38^, central-peripheral mixture (Guillain Barre syndrome) ^39^, enteric, or peripheral nervous systems dysfunction (anosmia, ageusia, myoclonus ^40^, neuropathic pain, and myalgia) ^41^. The increase in hospital admission and mortality rates in PD and other chronic neurological diseases during the COVID-19 pandemic is of imminent concern as the long-term sequelae are currently undetermined. Most neurological diseases, including PD, are dose-dependent on prescribed medications for symptomatic management. With the compounded neuropsychiatric symptoms due to social isolation and outpatient clinics, temporarily being suspended and irregular visits, these are causative in worsening PD symptoms among COVID-19 patients ^42^.

In this systematic review, we included studies with COVID-19 positive patients with a prior history of Parkinson’s disease. The most common manifestations due to COVID-19 in PD patients were found to be motor dysfunction. The majority of the studies showed a common motor deficit domain in the range of moderate to severe. Some of the patients presented with delirium, dementia, and encephalopathy among other COVID-19 complications. We posit that this may be related to the ACE2 mechanism in the nervous system. It is postulated that SARS-CoV-2 enters the cell, increases the activity of T cells, causing vasodilation, thrombosis, and hypoxemia, which may then lead to stroke and seizures. One theory supports the effect of the virus by stating that SARS-COV-2 attacks ACE 2 receptors in a multi-organ manner, targeting the brain. A study found that ACE2 receptors act locks on cells and the SARS-CoV-2 spike proteins act rapidly multiply on entering the cells. These ACE2 receptors control tissues in the eye, reproductive system, renal-excretory, digestive, and respiratory system, and 21 different regions of the brain, which require further deliberation in PD patients ^43^.

We found that the average age of all participants was 76.9 years with a male predominance, in addition to a prior disease duration of 11 years. The majority of the patients reported the presence of other comorbidities, of which the following were the most common: hypertension, diabetes, obesity, dyslipidemia, cardiovascular disease, immunocompromised, COPD, asthma, chronic renal and chronic liver diseases. Documented COVID-19 neurological complications include stroke, especially in younger patients, reflecting the hypercoagulable state, leukoencephalopathy, and hemorrhagic leukoencephalopathy. Although direct encephalitis was documented, it is rare but does occur. Further, cranial neuropathies are also infrequent but include the loss of sense of smell (cranial nerve 1) and Bell’s palsy (cranial nerve 7) ^44^. There have been neurocognitive signs, particularly in older age groups that warrant further associations to PD symptoms, clinical care, and management in the short and long term.

## 5. Limitations

We had certain limitations. The first included the presence of confounder factors. These related to the finding that multiple studies of patients with elderly PD patients had other comorbidities, possibly leading to an overrepresentation of findings. Secondly, the duration of PD and the severity of the pre-pandemic symptoms possibly influenced the synthesis of clinical outcomes due to COVID-19 infection. Thirdly, pertinent information, including prior and current medications and compliance to treatment was lacking, leading to a lack of generalized recommendations for PD and COVID-19 clinical care. While the inclusion of case reports may deviate from generalizations in our findings, we believe they are pertinent for long-term PD patients with acquired COVID-19 infection.

## 6. Conclusion

COVID-19 has a peaked mortality rate in older age groups, compounded with comorbid conditions. The CNS is increasingly susceptible to SARS-CoV-2, deteriorating neurological findings, particularly in patients with PD. We find that motor dysfunction, delirium, dementia, severe spasms of arms and legs, and encephalopathy are pertinent clinical findings in PD patients and ought to be addressed in current care and practice. The susceptible groups of COVID-19 positive patients with a history of PD are elderly males with 10 years or longer duration of illness and other comorbid conditions. It is still unclear how SARS-CoV2 affects the long-term health of the nervous system in PD patients. It is recommended that healthcare practitioners address diminished neurological outcomes, changes in hospital stay, and raised mortality in PD patients.

## Data Availability

ALL THE DATA ARE FULLY AVAILABLE IN THE MANUSCRIPT

## Acknowledgments

We would like to acknowledge Samia Jahan, Madiha Zaidi, Wanessa Matos, Sana Javed, Asma Mohammadi, and Sujan Poudel for their early contributions. All authors are thankful to Jack Michel, and the Larkin Health System for boosting scholarly research pursuits.

